# A Simple Score to Predict New-onset Atrial Fibrillation After Ablation of Typical Atrial Flutter

**DOI:** 10.1101/2023.05.18.23290204

**Authors:** Zhoushan Gu, Jincheng Jiao, Xiangwei Ding, Chao Zhu, Mingfang Li, Hongwu Chen, Weizhu Ju, Kai Gu, Gang Yang, Hailei Liu, Pipin Kojodjojo, Minglong Chen

**Author notes:** The first 3 authors contributed equally to this work. Corresponding author: Hailei Liu, MD, Division of Cardiology, The First Affiliated Hospital of Nanjing Medical University. 300#, Guangzhou Road, Nanjing, 210029, P.R. China.

## Abstract

**Background:** New-onset atrial fibrillation (NeAF) is common after cavotricuspid isthmus-dependent counterclockwise atrial flutter (CCW-AFL) ablation. This study aimed to investigate a simple predictive model of NeAF after CCW-AFL ablation.

**Methods and Results:** From January 2013 to December 2017, consecutive patients receiving CCW-AFL ablation were enrolled from three centers. Clinical, echocardiographic, and electrocardiographic data were collected and followed. Patients from two centers and another center were assigned into the derivation and validation cohorts, respectively. In the derivation cohort, logistic regression was performed to evaluate the ability of parameters to discriminate those with and without NeAF. A score system was developed and then validated. Two hundred seventy-one patients (mean 59.7±13.6 age; 205 male) were analyzed. During follow-up (73.0±6.5 months), 107 patients (39.5%) had NeAF. 190 and 81 patients were detected in the derivation and validation cohorts, respectively. Hypertension, age ≥70 years, left atrial diameter ≥42 mm, P wave duration ≥120 ms and the negative component of flutter wave in lead II ≥120 ms were selected as the final parameters. A weighted score was used to develop the HAD-AF score ranging from 0 to 9. In the derivation cohort, area under the receiver operating characteristic curve (AUC) was 0.938 (95% CI 0.902-0.974), superior to those of currently used CHA2DS2-VAS_C_ (0.679, 95% CI 0.600-0.757) and HATCH scores (0.651, 95% CI 0.571-0.730) (P<0.001). Performance maintained in the validation cohort.

**Conclusions:** 39.5% of patients developed NeAF in 6 years after CCW-AFL ablation. HAD-AF score can reliably identify patients likely to develop NeAF after CCW-AFL ablation.

**Clinical Perspective:** *What Is New?:*

1. During a follow-up period of more than 6 years after CCW-AFL ablation, 107 of 271 (39.5%) patients developed NeAF.
2. HAD-AF score, based on easily obtainable clinical, echocardiographic and electrocardiographic parameters, could better predict development of NeAF after CCW-AFL ablation (area under the receiver operating characteristics curve [AUC], 0.938), compared with currently used HATCH score (AUC, 0.651) and CHA2DS2-VAS_C_ score (AUC, 0.679) (P<0.001).

*What Are the Clinical Implications?:* In CCW-AFL patients with a HAD-AF score >4, close postoperative follow-up for earlier detection of AF should be recommended, or the option of concomitant AF ablation could be considered during the shared decision-making process.

---

Typical cavotricuspid isthmus-dependent counterclockwise atrial flutter (CCW-AFL) is a common clinical arrhythmia. Currently, radiofrequency catheter ablation (RFCA) is widely performed to treat CCW-AFL, given its high success rate and low risk. However, even without a history of atrial fibrillation (AF), such patients with a successful CCW-AFL ablation have significantly higher risk of new-onset AF (NeAF) compared to the general population, making them more susceptible to stroke and heart failure.^1-5^ For patients at risk of NeAF, close postoperative follow-up for earlier detection of AF, or even performing concomitant ablation of AF and CCW-AFL could be potentially desirable.^6^ In this context, accurate identification of patients most likely to develop NeAF is of great clinical relevance in terms of pre-procedural counselling and shared decision making with the patient.

At present, the reported clinical predictors of NeAF include age, obesity and renal insufficiency, etc., but the clinical applicability of univariate predictors is limited.^7-9^ Thus, some studies have proposed score systems such as HATCH and CHA2DS2-VASc scores based on clinical parameters in predicting NeAF after CCW-AFL ablation. Nevertheless, the predictive performance of these scores remained unsatisfactory.^10, 11^ Apart from clinical parameters, left atrial remodeling indicators are also closely associated with AF.^12^ Currently, several studies found that the indicators of both left atrial structural and electrical remodeling derived from the echocardiogram and electrocardiogram (ECG) had independent predictive value beyond clinical parameters.^5, 13, 14^ We hypothesized that a score system that combines clinical parameters with the readily available echocardiographic and ECG parameters is more effective in predicting NeAF post CCW-AFL ablation than the currently available risk scoring algorithms.

In this multicenter prospective cohort study, we aimed to investigate the long-term risk of NeAF after CCW-AFL ablation, to develop such a scoring system to better estimate future risk of NeAF, and to compare its performance with pre-existing scoring systems.

## Methods

### Patient population

This is a multicenter prospective cohort study. From January 2013 to December 2017, participants from the following three Chinese centers were consecutively enrolled: The First Affiliated Hospital of Nanjing Medical University, Affiliated Hospital of Nantong University, and Jiangsu Taizhou People’s Hospital. The inclusion criteria were as follows: (1) aged ≥18 years; (2) receiving CCW-AFL catheter ablation. Exclusion criteria were as follows: (1) any documented AF episodes before ablation; (2) abnormal thyroid function; (3) previous cardiac surgery history; (4) previous ablation of atrial tachycardia or atrial flutter; (5) life expectancy <12 months; (6) refused to participate. Informed consents were obtained and the study protocols were approved by the Ethics Committees of the above three centers.

### Clinical data collection and ECG parameters measurement

Before the index procedure, patients’ baseline data were obtained, including gender, age, BMI, comorbidities, echocardiography, and 12-lead ECG both under sinus rhythm (SR) and CCW-AFL (25 mm/s, 10 mm/mV). If ECG under SR or CCW-AFL was unobtainable after admission, the most recent corresponding ECG free from any antiarrhythmic drugs (AADs) influence (administration of AADs within 1 month before ECG acquisition) was collected. The ECG parameters reflecting electrical activity of the left atrium under SR and CCW-AFL, associated with atrial electrical remodeling as proven by our previous study,^15^ were measured by two independent cardiologists to provide an average value of three different beats using Adobe Acrobat XI Pro (Adobe, California, USA) (Figure S1 in the Data Supplement). These included: (1) duration of the negative component of flutter wave in lead II (D_FNII_); (2) amplitude of the negative component of flutter wave in lead II (A_FNII_); (3) amplitude of negative component of the P wave in lead V1 under SR (A_PNV1_); (4) duration of negative component of the P wave in lead V1 under SR (D_PNV1_); and (5) P wave duration in lead II under SR (D_PII_). The values obtained from the two cardiologists were averaged. Then, P wave terminal force in Lead V1 (P_tfV1_) under SR was calculated using the formula A_PNV1_*D_PNV1_ (mm*ms).

### Electrophysiological study and catheter ablation

Before the procedure, AADs were discontinued for at least 5 half-lives. Oral anticoagulants were administered for at least three weeks prior to ablation. Transesophageal echocardiography was performed to exclude intra-cardiac thrombosis before ablation. Under routine local anesthesia, a quadripolar and a decapolar catheter were introduced to the right ventricle and coronary sinus via femoral approach, respectively. Activation mapping using the three-dimensional mapping system (Carto, Ensite, or Rhythmia) and entrainment mapping were applied to confirm the diagnosis of CCW-AFL. Subsequently, linear ablation between the tricuspid annulus and the inferior vena cava was performed with irrigated catheters under the power setting of 35-40 W and flow rate of 17-30 ml/min. Bidirectional block across the cavotricuspid isthmus line was defined as procedural endpoint.

### Follow-up of patients

AADs were discontinued after a successful ablation. Anticoagulation was continued for at least 3 months postoperatively and discontinued at the physicians’ discretion, depending on the occurrence of NeAF and CHA2DS2-VASc score. Patients were followed up every three months within the first year, and every six months thereafter. ECG and 24-hour Holter monitoring were performed during each follow-up visit. Additional ECGs and 24-hour Holter monitoring were obtained in the event of arrhythmic symptoms. CCW-AFL recurrence was defined as documented CCW-AFL episode lasting >30s after a 3-month blanking period postoperatively. NeAF was defined as any AF episode longer than 30s postoperatively.^16^

### Statistical analysis

Continuous variables were described as mean ± standard derivation (SD). Unpaired Student’s t-test was used in normally distributed variables. Friedman test was used in non-normally distributed variables. Categorical variables were presented as counts with percentages and compared using Chi-square tests or Fisher’s exact tests. Two-sided *P*<0.05 was considered statistically significant.

Patients from two centers were assigned into derivation cohort, and patients from the other center were assigned into validation cohort. A score system was derived from the derivation cohort as follows. All patients in the derivation cohort were assigned to new-AF or non-AF groups according to the presence or absence of NeAF during follow-up. Univariate and multivariate logistic regression analyses were conducted to identify the predictors of NeAF. For ease of clinical use, continuous variables that were statistically different were dichotomized with ROC curves to identify optimal cut-points for discrimination, which were rounded to the nearest clinically significant integer when applicable. Variables that were significant in univariate analysis were included into the model, with attention paid to avoid clinically relevant collinearity. A prediction score was created with the variables and strength of association by β coefficients rounded to the nearest integer.^17^

Subsequently, its diagnostic performance was evaluated with the area under the receiver operating characteristics curve (AUC) values. To evaluate incremental discrimination beyond existing criteria, we used Delong’s test to compare the AUC values from our derived score system with those from HATCH and CHA2DS2-VASc score systems. The Hosmer-Lemeshow goodness of fit test (P≥0.05) was evaluated to access calibration for model validation. The diagnostic performance of this score system was validated in a separate validation cohort. All statistical analyses were performed using SPSS software version 26.0.

## Results

### Baseline characteristics and follow-up results

A total of 424 patients were assessed for eligibility, and 145 were excluded. Eight patients (2.9%) were lost to follow-up. Thus, 271 patients (mean age, 59.7±13.6 years; 205 male) were included in the final analysis (Figure 1). Ablation procedural endpoint was reached in all subjects. Major periprocedural complications were observed in 5 patients (1.8%), all of whom had vascular access complications. During a follow-up period of 73.0±6.5 months, 11 patients (4.1%) had CCW-AFL recurrence and underwent a second ablation. 107 cases (39.5%) had NeAF, with 48, 37 and 22 cases in the first year, first to third years, and after 3 years after ablation, respectively. Additionally, due to new-onset symptomatic AF, 57 cases underwent AF ablation and 48 were prescribed with AADs. Patients from the First Affiliated Hospital of Nanjing Medical University and Affiliated Hospital of Nantong University were assigned into derivation cohort (n=190, 70.1%), and patients from Jiangsu Taizhou People’s Hospital were assigned into validation cohort (n=81, 29.9%), which shared similar clinical characteristics (Table 1).

**Figure 1.**
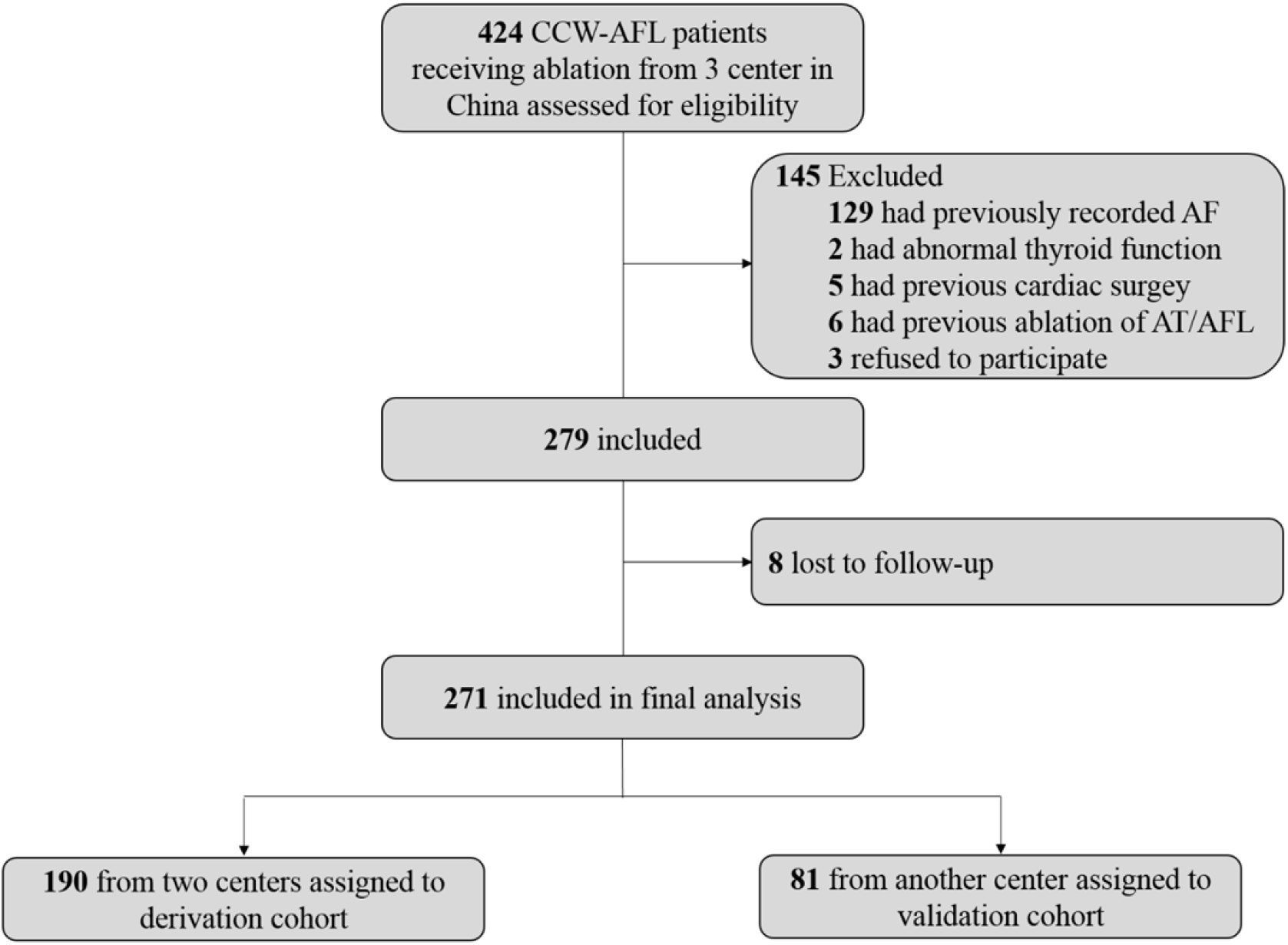
The algorithm of patient enrollment. AF indicates atrial fibrillation; AFL, atrial flutter; AT, atrial tachycardia; CCW-AFL, counterclockwise atrial flutter.

**Table 1.** Baseline characteristics of patients in derivation and validation cohorts.

|  | All<br>(n=271) | Derivation cohort<br>(n=190) | Validation cohort<br>(n=81) | *P*<br>value |
| --- | --- | --- | --- | --- |
| Age, y | 59.3±13.8 | 58.4±14.9 | 61.4±10.4 | 0.107 |
| Male, n (%) | 205 (75.6) | 147 (77.4) | 58 (71.6) | 0.313 |
| BMI | 24.8±2.7 | 24.7±2.5 | 25.1±3.1 | 0.280 |
| Comorbidities, n (%) |  |  |  |  |
| Hypertension | 124 (45.8) | 85 (44.7) | 39 (48.1) | 0.606 |
| DM | 64 (23.6) | 51 (26.8) | 13 (16.0) | 0.058 |
| COPD | 9 (3.3) | 6 (3.2) | 3 (3.7) | 0.819 |
| Heart failure | 41 (15.1) | 28 (14.7) | 13 (16.0) | 0.783 |
| Stroke/TIA | 26 (9.6) | 19 (10.0) | 7 (8.6) | 0.728 |
| CHD | 33 (12.2) | 23 (13.2) | 10 (12.3) | 0.956 |
| eGFR <60 ml/min | 22 (8.1) | 17 (8.9) | 5 (6.2) | 0.446 |
| CHA2DS2-VASc score | 2.0±1.5 | 2.0±1.4 | 1.9±1.5 | 0.898 |
| HAS-BLED score | 1.0±1.0 | 1.0±1.0 | 0.7±0.9 | 0.059 |
| HATCH score | 1.1±1.3 | 1.1±1.2 | 1.1±1.5 | 0.948 |
| Echocardiography |  |  |  |  |
| LAD, mm | 39.6±5.5 | 39.3±5.7 | 40.2±5.2 | 0.239 |
| LVEF, % | 59.0±7.8 | 59.5±7.4 | 57.9±8.8 | 0.121 |
| ECG |  |  |  |  |
| PR interval, ms | 176.7±32.8 | 179.9±35.1 | 175.2±25.1 | 0.278 |
| D<sub>FNII</sub>, ms | 112.0±30.2 | 109.7±30.1 | 113.4±23.3 | 0.322 |
| TCL, ms | 210.3±17.2 | 210.6±19.1 | 209.7±11.7 | 0.678 |
| A<sub>FNII</sub>, mV | 0.20±0.05 | 0.20±0.05 | 0.21±0.04 | 0.131 |
| $A_{PNV1}$, mV | $0.06 \pm 0.05$ | $0.06 \pm 0.05$ | $0.06 \pm 0.04$ | 0.249 |
| $D_{PNV1}$, ms | $34.5 \pm 28.9$ | $35.4 \pm 31.3$ | $32.1 \pm 22.4$ | 0.390 |
| PtfV1, mm*ms | $2.4 \pm 2.8$ | $2.4 \pm 2.9$ | $2.4 \pm 2.6$ | 0.968 |
| $D_{PII}$, ms | $116.0 \pm 11.7$ | $116.3 \pm 11.0$ | $115.0 \pm 13.4$ | 0.401 |

Values are presented as mean ± SD or n (%). $A_{FNII}$ indicates amplitude of the negative component of flutter wave in lead II; $A_{PNV1}$ , amplitude of negative component of the P wave in lead V1 under sinus rhythm; BMI, body mass index; CHD, coronary heart disease; COPD, Chronic obstructive pulmonary disease; $D_{FNII}$ , duration of the negative component of flutter wave in lead II; DM, diabetes mellitus; $D_{PII}$ , P wave duration in lead II under sinus rhythm; $D_{PNV1}$ , duration of negative component of the P wave in lead V1 under sinus rhythm; ECG, electrocardiogram; LAD, left atrial diameter; LVEF, left ventricular ejection fraction; $P_{tfV1}$ , P wave terminal force in Lead V1; TCL, total cycle length; and TIA, transient ischemic attack.

### Univariate and multivariate analyses

In the derivation cohort of 190 patients, 76 (40.0%) had NeAF. Statistical significant differences were found between the groups with and without NeAF in terms of age, hypertension, CHA2DS2-VASc score, HATCH score, left atrial diameter (LAD), D_FNII_ and D_PII_, based on univariate analysis (Table 2). For clinical ease of use, we converted the continuous variables, such as age, LAD, D_FNII_, and D_PII_ into categorical variables (Table S1 in the Data Supplement). By conducting a multivariate analysis on these individual parameters with a predictive value, it was found that hypertension, age ≥70 years, LAD ≥42 mm, D_PII_ ≥120 ms, and D_FNII_ ≥120 ms were independent predictors of NeAF (Table 3).

**Table 2.**
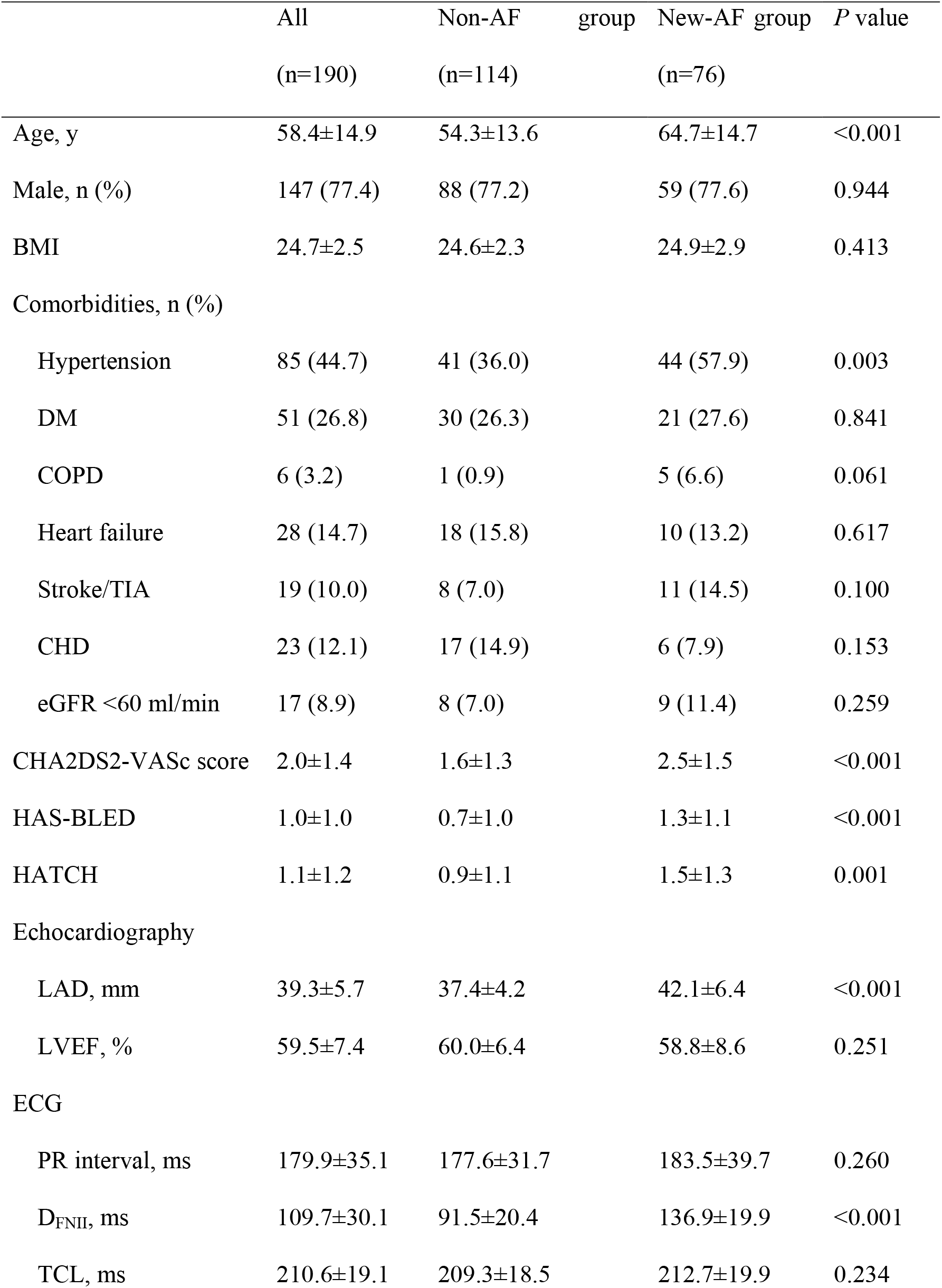
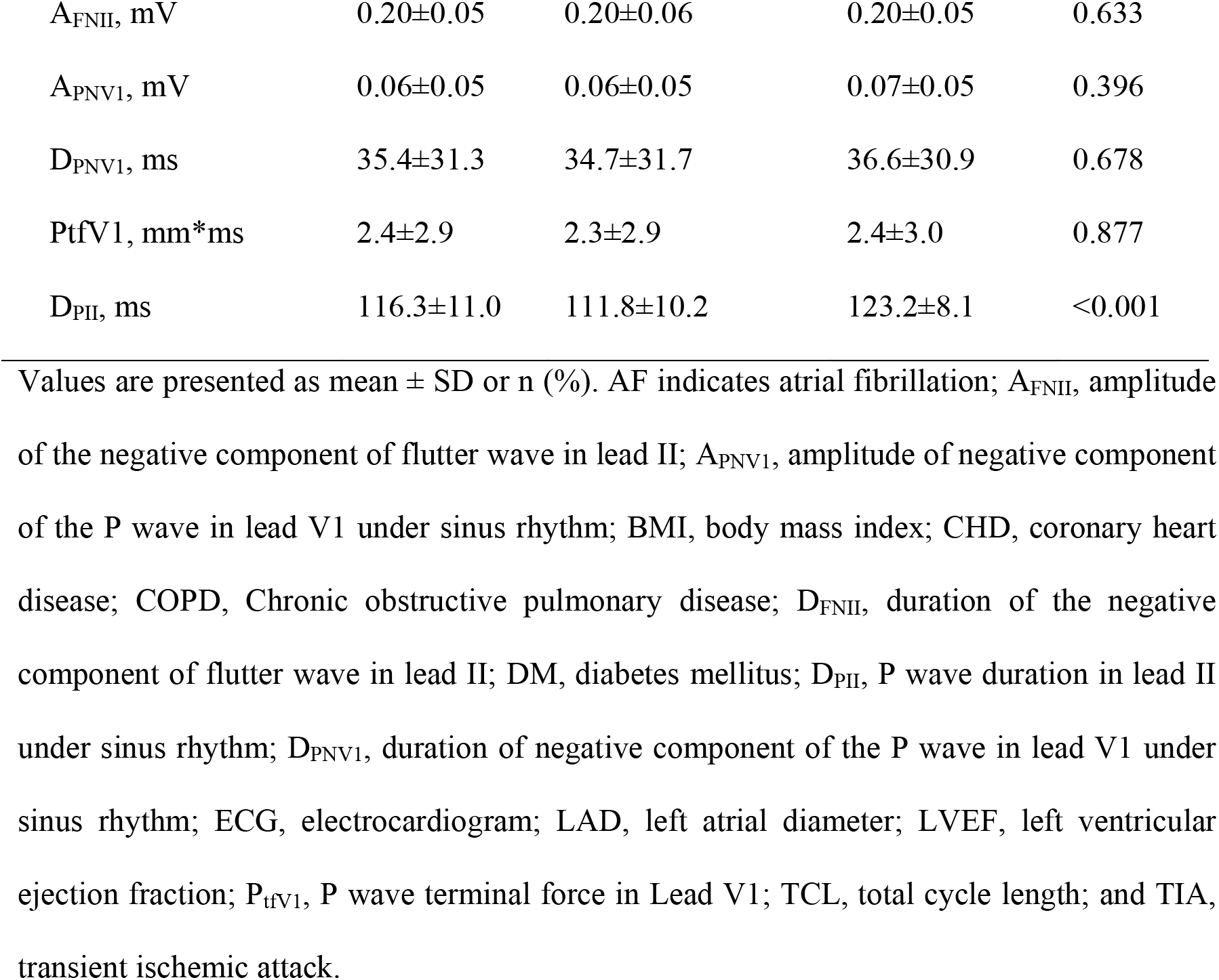
Comparison of the parameters between patients with and without new-onset AF in the derivation cohort.

**Table 3.**
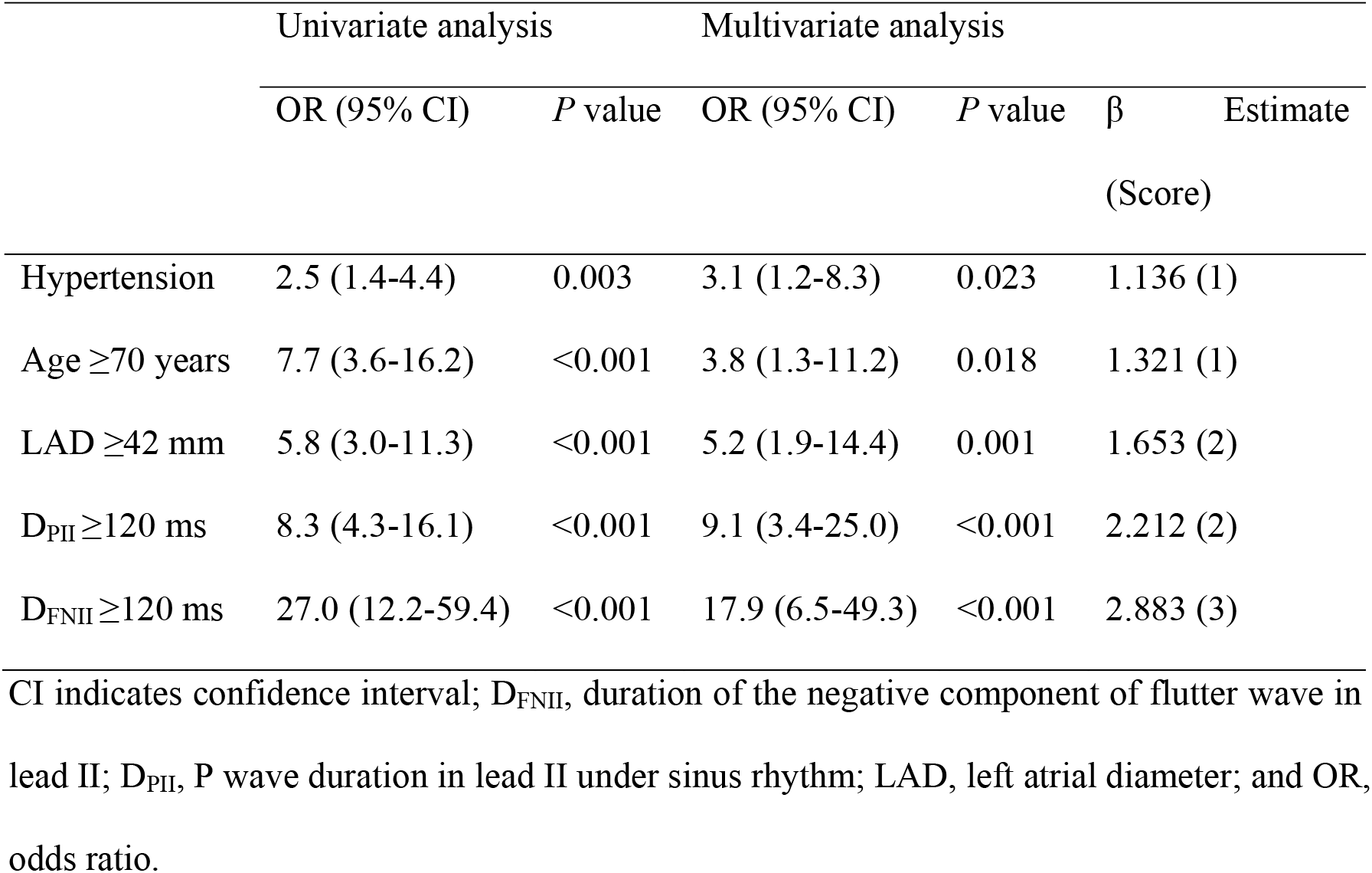
Univariate and multivariate Cox regression analysis for identifying predictors of new-onset atrial fibrillation.

### Derivation of the score system

The HAD-AF score, ranging from 0 to 9, was developed to predict NeAF based on the β coefficients of the above 5 variables (Table 3), with one point for hypertension and age ≥70 years, two points for LAD ≥42 mm and D_PII_ ≥120 ms, and three points for D_FNII_ ≥120 ms (Figure 2). The probability of NeAF increased along with HAD-AF score (Figure 2). Model-based probabilities closely matched the observed prevalence for each given score value, indicating good calibration (Figure 3). The AUC values of HAD-AF, HATCH and CHA2DS2-VASc score systems were 0.938 (95% CI 0.902-0.974), 0.651 (95% CI 0.571-0.730), and 0.679 (95% CI 0.600-0.757) (P<0.001), respectively (Figure 4, panel A). Additionally, a HAD-AF score >4 could predict NeAF with a sensitivity of 0.829 and a specificity of 0.939.

**Figure 2.**
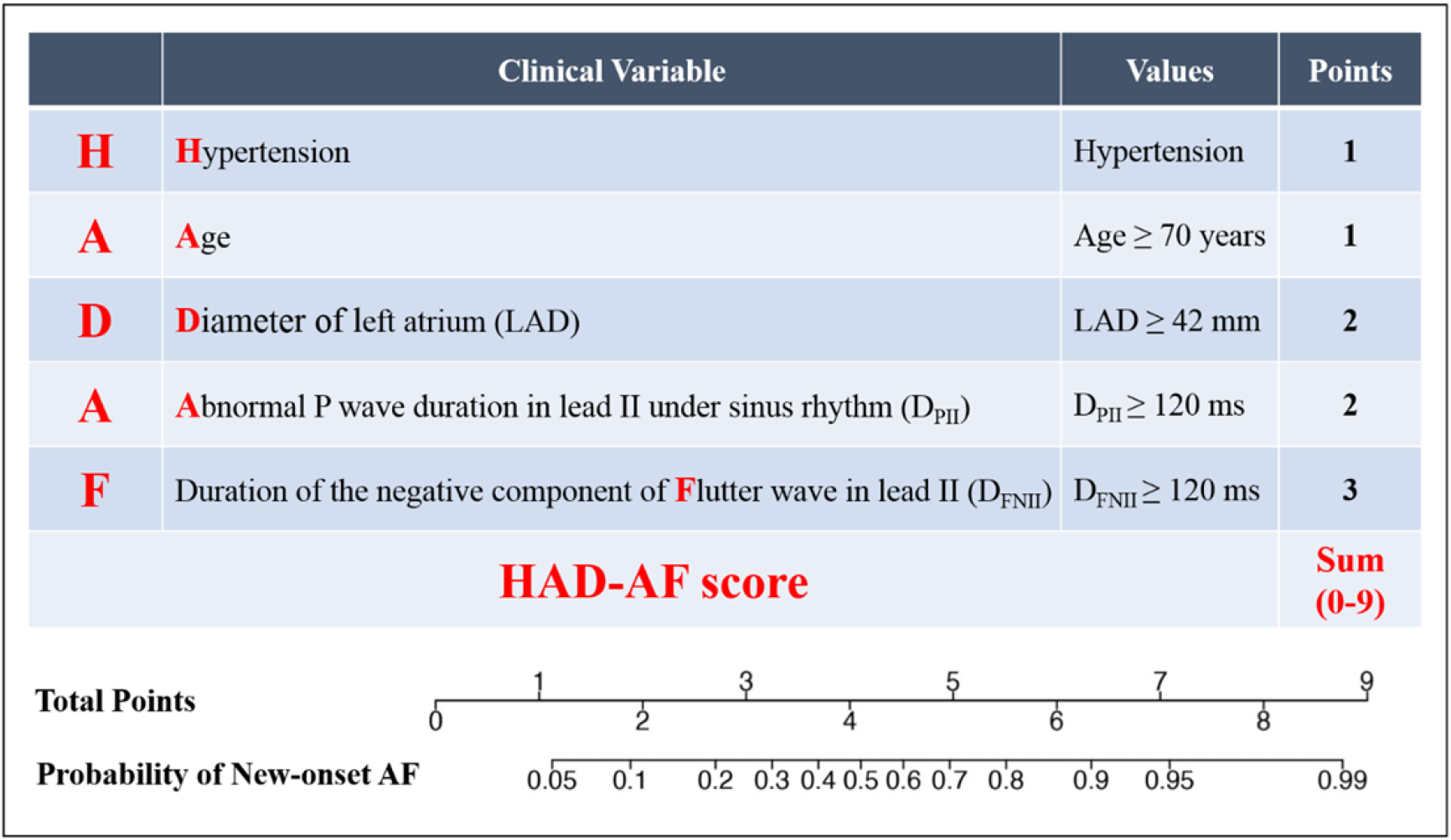
Description of the HAD-AF score. Description of the HAD-AF score and point allocations for each clinical characteristic (top), with associated probability of development of new-onset atrial fibrillation based on the total score as estimated from the model (bottom).

**Figure 3.**
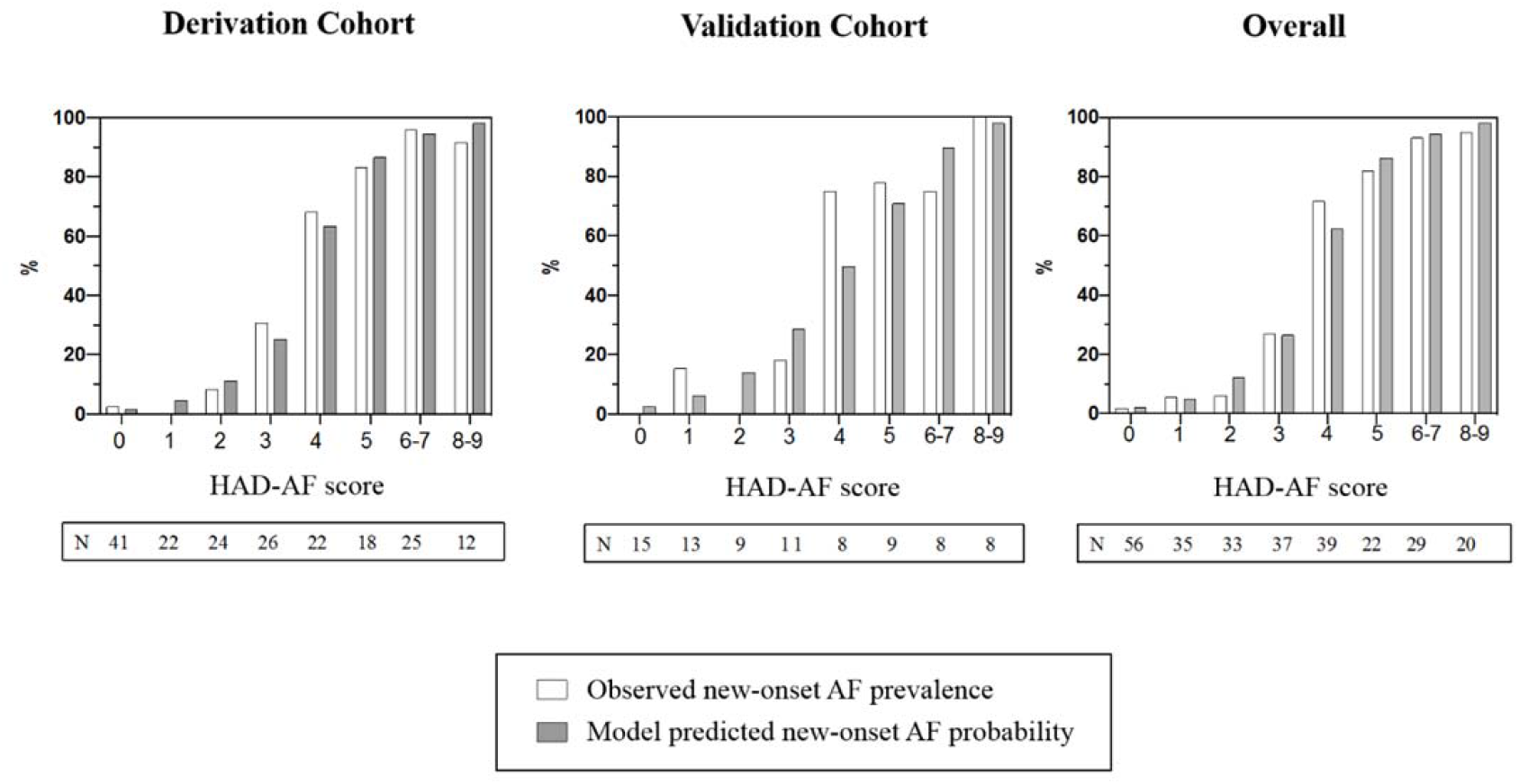
Calibration of the HAD-AF score. The Hosmer-Lemeshow goodness-of-fit test results using deciles of predicted probabilities were P=0.518, 0.199, and 0.640 for the derivation, validation, and overall sample, respectively, indicating support for a properly calibrated model. AF indicates atrial fibrillation.

**Figure 4.**
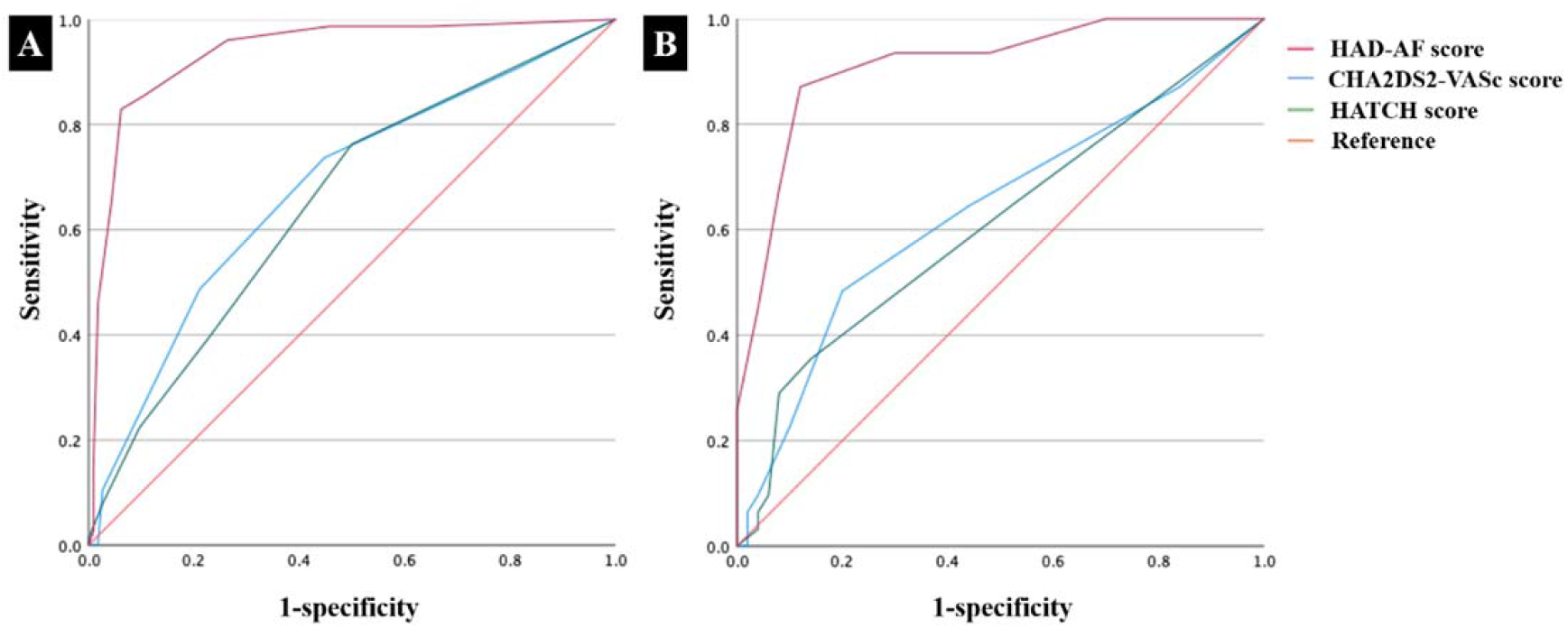
Prediction value of different score systems in the derivation and validation cohorts. Panel A, in the derivation cohort, AUC= 0.938, 0.651, and 0.679 for HAD-AF, HATCH, and CHA2DS2-VASc score systems, respectively. Panel B, in the validation cohort, AUC= 0.912, 0.610, and 0.635 for HAD-AF, HATCH, and CHA2DS2-VASc score systems, respectively. AUC indicates area under the receiver operating characteristic curve.

### Validation of HAD-AF score

Among 81 patients assigned to the validation cohort, 31 (38.3%) experienced NeAF. Results of univariate regression analysis in the validation cohort were similar to those in the derivation cohort (Table S2 in the Data supplement). Model-based probabilities remained closely matched with the observed prevalence for each given score value (Figure 3). The AUC of the HAD-AF score to predict NeAF was 0.912 (95% CI 0.846-0.978), higher than that of the HATCH (0.610, 95% CI 0.480-0.741) and CHA2DS2-VAS_C_ score systems (0.635, 95% CI 0.506-0.764) (P<0.001), validating a higher predictive value of HAD-AF score (Figure 4, panel B).

## Discussion

The major findings of this multicenter prospective cohort study were as follows: (1) in the 6 years after a successful CCW-AFL ablation, 39.5% of patients developed NeAF; (2) Hypertension, age ≥70 years, LAD ≥42 mm, D_FNII_ ≥120 ms, and D_PII_ ≥120 ms were independent predictors of NeAF; (3) the HAD-AF score based on the parameters of hypertension, age, LAD, D_FNII_, and D_PII_ exhibited a higher predictive value than the currently used HATCH and CHA2DS2-VASc score systems.

CCW-AFL and AF are two different forms of atrial arrhythmias sharing similar risk factors, pathophysiological and electrophysiological mechanisms, which could occur concomitantly or separately. In CCW-AFL patients with concomitant AF episodes, combined ablation of CCW-AFL and AF was proved to be beneficial, given a high recurrence rate of AF post CCW-AFL ablation alone.^18-20^ However, as shown in this study, even in CCW-AFL patients without previously recorded AF, the NeAF incidence could reach as high as 39.5% during a median follow-up period of 73.0 months. Additionally, these patients with NeAF were usually accompanied with comorbidities (with an average CHA2DS2-VASc score of 2.5±1.5 as shown in this study), which made them more susceptible to major adverse clinical events once AF ensues.^21^ Thus, intensive follow-up is necessary, and patients should be counselled pre-procedurally for consideration of concomitant AF ablation, at the expense of a longer procedure and a small increase in procedural risks, compared to CCW-AFL ablation alone.^19^

Atrial remodeling is strongly linked to the initiation and maintenance of AF.^12, 22-24^ Clinical parameters, such as age, hypertension and heart failure etc. are clinical predictors of AF, most likely caused by increasing left atrial stretch, and promoting structural changes and adverse atrial remodeling. Previous studies found that score systems based on clinical parameters like CHA2DS2-VASc and HATCH scores have modest ability to predict NeAF following CCW-AFL ablation.^10, 11^ However, clinical variables chosen in these scores are surrogate markers for atrial remodeling and therefore have diminished predictive value. In our study, AUC values of CHA2DS2-VASc and HATCH scores in predicting NeAF following CCW-AFL ablation were only 0.679 and 0.651, respectively, which were similar to previous studies.^10, 11^

Left atrial remodeling is usually characterized by alterations in left atrial structure and/or electrical activity,^22^ manifested as increased LAD or left atrium volume, slowed wavefront propagation and refractoriness.^25^ In this study, increased LAD was associated with a greater risk of NeAF after CCW-AFL RFCA, consistent with other studies.^14^ The current study also showed that the ECG parameters reflecting left atrial activation time, namely D_PII_, D_FNII_, were also associated with NeAF, consistent with our previous study with one-year follow-up.^15^

The HAD-AF score, based on the clinical and left atrial remodeling-related indicators, exhibited a higher value in predicting NeAF after CCW-AFL ablation (AUC=0.938), compared with CHA2DS2-VASc and HATCH scores. In CCW-AFL patients with a HAD-AF score >4, close postoperative follow-up for earlier detection of AF should be recommended, or the option of concomitant AF ablation could be considered during the shared decision-making process.

### Study limitation

Firstly, due to practical constraints^26^ and early initiation of this study, the absence of long term Holter monitoring and implanted loop recorder might lower the detection rate of preoperative and postoperative AF. However, the preoperative management and postoperative follow-up flow in this study were still considered as a standardized screening protocol in such patients.^27^ Secondly, atrial fibrosis could also be assessed by cardiac magnetic resonance imaging or possibly biomarkers. However, to simplify the HAD-AF risk score, magnetic resonance imaging parameters or biomarkers were not routinely measured in this study. Thirdly, as AF episode of over 30s was set as the endpoint, a larger multicenter prospective study was needed to test the value of HAD-AF score to improve clinical outcomes.

## Conclusions

39.5% of patients develop NeAF 6 years after CCW-AFL ablation. HAD-AF score, based on easily accessible clinical, echocardiographic and ECG parameters, can more effectively predict NeAF after CCW-AFL ablation and better guide pre-procedural consent and counselling, compared with currently used HATCH score and CHA2DS2-VASc score.

## Data Availability

All data referred to in the manuscript is available if needed.

## Nonstandard Abbreviations and Acronyms

AADs: antiarrhythmic drugs
A_FNII_: amplitude of the negative component of flutter wave in lead II
A_PNV1_: amplitude of negative component of the P wave in lead V1 under sinus rhythm
CCW-AFL: counterclockwise atrial flutter
D_FNII_: duration of the negative component of flutter wave in lead II
D_PII_: P wave duration in lead II under sinus rhythm
D_PNV1_: duration of negative component of the P wave in lead V1 under sinus rhythm
LAD: left atrial diameter
LVEF: left ventricular ejection fraction
NeAF: new-onset atrial fibrillation

## Acknowledgments

None.

## Disclosures

None.

## Supplemental materials

Supplemental Tables S1-S2

Supplemental Figure S1

## Notes

**Conflict of interest statement:** none.

### Competing Interest Statement

The authors have declared no competing interest.

### Author Declarations

The Ethics Committees of the First Affiliated Hospital of Nanjing Medical University, Affiliated Hospital of Nantong University, and Jiangsu Taizhou People's Hospital.

